# Interference of urinary albumin-to-creatinine ratio measurement by glycosuria: clinical implications when using SGLT-2 inhibitors

**DOI:** 10.1101/2022.09.16.22280029

**Authors:** D Chapman, PK Judge, RJ Sardell, N Staplin, T Arnold, D Zhu, S Ng, S Moffat, MJ Landray, C Baigent, M Hill, R Haynes, S Clark, WG Herrington

## Abstract

Albuminuria is used for chronic kidney disease (CKD) screening, diagnosis, staging, and monitoring. A change in albuminuria has been proposed as a surrogate outcome for CKD progression. High glucose concentration interferes with Jaffe serum creatinine assays but the extent to which glycosuria biases measurement of urinary albumin-to-creatinine ratio (uACR) is uncertain. Any interference would have implications as the use of sodium-glucose co-transporter-2 (SGLT-2) inhibitors increases. We performed laboratory-based interference studies on urine samples from 333 adults with CKD stages 3-4. Samples were separated into four aliquots: a reference aliquot and three aliquots spiked with increasing concentrations of glucose solution representing the range expected in patients taking SGLT-2 inhibitors (28, 111 and 333 mmol/L). uACR was assayed using Jaffe and enzymatic methods. Median (Q1-Q3) uACR in reference samples was 63 (17-150) mg/mmol. Glucose spiking did not interfere with uACR estimation using enzymatic creatinine assays. For the Jaffe assay, the presence of 28 mmol/L of glucose resulted in a -1.5% mean bias in uACR (95% confidence interval -1.9 to -1.1%) which increased to a -2.5% bias (−3.2 to -1.9%) at a concentration of 333 mmol/L. Overestimation of urinary creatinine concentration increased substantially with decreasing creatinine concentration (i.e. dilute urine). In this cohort, interference of the Jaffe assay by glucose spiking resulted in 2-5% of uACR samples having a ≥10% spurious reduction in uACR (on its original scale). Given the increasing use of SGLT-2 inhibitors, we suggest uACR measured using Jaffe creatinine assays should be avoided.

## Introduction

Urinary albumin-to-creatinine ratio (uACR) measured on spot samples provides a convenient method to screen for, diagnose, stage and monitor chronic kidney disease (CKD),^1,2^ and change in albuminuria is proposed as a surrogate outcome for CKD progression in clinical trials.^3,4^ Sodium-glucose co-transporter-2 (SGLT-2) inhibitors increase urinary glucose excretion to 50-80g/day under normoglycaemic and modest hyperglycaemic conditions, and >100g/day in people with diabetes and hyperfiltration.^5^ This can equate to urinary glucose concentrations as high as 500mmol/L. SGLT-2 inhibitors are increasingly used due to important beneficial effects on risk of cardiovascular disease and CKD progression.^6-12^

High serum glucose is known to interfere with the Jaffe reaction used to measure serum creatinine,^13^ with bias exceeding 10% with glucose concentrations >20 mmol/L in some assays.^14^ This problem can be circumvented by use of enzymatic creatinine assays,^15^ but such assays are more expensive and use is low even in high-income countries (only about one-half of the UK’s reference laboratories use enzymatic methods).^16^ Like serum, urinary glucose may interfere with urinary creatinine measurement, and the potentially high levels of glycosuria resulting from use of SGLT-2 inhibitors could have important clinical implications.^17-21^ However, any bias of uACR measurements when using different assays is unknown. We aimed to address this uncertainty using laboratory interference studies and urine samples from patients with CKD spiked with a range of glucose concentrations intended to represent the range expected in patients taking SGLT-2 inhibitors.

## Methods

Early morning urine samples collected and stored as part of the UK Heart and Renal Protection (HARP)-III trial (previously reported)^22^ were used for laboratory interference experiments. UK HARP-III randomized 414 participants ≥18 years with CKD stages 3/4 to irbesartan or sacubitril/valsartan. Urine samples have been stored at -80°C, a temperature reported to maintain stability of uACR measurement, prior to analysis.^23,24^ 370 participants’ samples with a urine volume of at least 1mL were available for this study.

### Laboratory methods

Each urine sample was thawed, mixed by inversion, and samples separated into four 245µl aliquots. One aliquot had 70µl of deionised water added (reference sample), the remaining aliquots were spiked with 70µl of either 125, 500 or 1500 mmol/L glucose solution (produced by diluting Merck product: G8769 in deionised water). This provided a final glucose concentration of 28, 111 and 333 mmol/L, respectively (the range expected in patients on SGLT-2 inhibitors or with poorly controlled diabetes).^25^

Urine albumin was measured by immunoturbimetric method, urine creatinine was measured using both Jaffe and enzymatic methods traceable to IDMS calibration standards, and urinary glucose was measured using enzymatic method, on a Beckman Coulter DxC700AU Clinical Chemistry Analyser (Beckman Coulter, Inc., Brea CA) using the manufacturer’s reagents and protocols. The NDPH Wolfson is a UKAS accredited testing laboratory No 2799 and these assays were on their ISO/IEC 17025 Schedule of Accreditation at the time of this study. The repeatability coefficient of variation, determined using at least two concentration levels of each measurement, was <1.5% for all methods.

### Statistical analyses

Analyses used Bland Altman plots comparing the mean and difference of albumin, creatinine and uACR measurements. Reference and spiked samples were obtained for each level of glucose spiking.^26^ For assessments of bias, albumin and uACR measurements were log-transformed prior to plotting. Where the slope between the mean and difference of the samples was significantly different from zero (i.e. the bias between reference and spiked samples was not constant), a regression line was fitted. The difference in log uACR before and after the addition of glucose was also plotted against mean creatinine, both overall and by subgroups of uACR at randomization. For samples with uACR below the measureable range, the lower limit of quantification was imputed. Analyses were performed using SAS version 9.4 and R version 4.1.2.

## Results

From the 370 participants’ urine samples, 37 (10%) samples with endogenous glycosuria ≥5.6 mmol/L were excluded leaving 333 participants’ samples for analyses. Median (IQR) uACR was 63 (17-150) mg/mmol and 30 (9%) had normoalbuminuria, 72 (22%) microalbuminuria and 231 (69%) macroalbuminuria (Table 1). Median (IQR) urinary glucose concentration before spiking with glucose was 0.33 (0.33-0.57) mmol/L.

**Table 1:** Baseline characteristics.

| **Variable** | **n=333** |
| --- | --- |
| Age |  |
| Mean (SD) | 62 (14) |
| <50y | 63 (19%) |
| ≥50y-<70y | 158 (47%) |
| ≥70y | 112 (34%) |
| Sex |  |
| Male | 245 (74%) |
| Female | 88 (26%) |
| Ethnicity |  |
| White | 303 (91%) |
| South Asian | 14 (4%) |
| Black | 7 (2%) |
| Other | 9 (3%) |
| Prior diabetes | 107 (32%) |
| **CKD-EPI estimated glomerular filtration rate at randomisation (mL/min/1.73m<sup>2</sup>)** |  |
| Mean (SD) | 35.4 (10.8) |
| <30 | 126 (38%) |
| ≥30 to <45 | 140 (42%) |
| ≥45 | 66 (20%) |
| Not available | 1 |
| **Urine albumin-to-creatinine ratio at randomisation (mg/mmol)** |  |
| Geometric mean (approx SE) | 43 (4) |
| Median (IQR) | 63 (17-150) |
| <3 | 30 (9%) |
| ≥3 to <30 | 72 (22%) |
| ≥30 | 231 (69%) |
| **Urine creatinine at randomisation (mmol/L)** |  |
| Geometric mean (approx SE) | 6.06 (0.16) |
| Median (IQR) | 6.18 (4.20-8.34) |
| <2.5 | 14 (4%) |
| ≥2.5 to <5 | 105 (32%) |
| ≥5 | 214 (64%) |
| **Glucose concentration before spiking (mmol/L)** |  |
| Median (IQR) | 0.33 (0.33-0.57) |

Values are n (%), mean (SD), geometric mean (~SE), or median (IQR).
CKD-EPI indicates Chronic Kidney Disease Epidemiology Collaboration; IQR, interquartile range.

### Interference studies assessed using log uACR

There was no evidence that spiking with glucose had any effect on urinary albumin measurements at 28 or 111 mmol/L glucose concentration, but a 0.5% bias emerged at 333 mmol/l (Supplemental Figure S1). There was no bias for enzymatic creatinine measurements (Figure 1A). Consequently, overall there was only a small bias in log uACR measurement when an enzymatic method was used and when urine glucose concentration was 333 mmol/L (Figure 2A).

**Figure 1:**
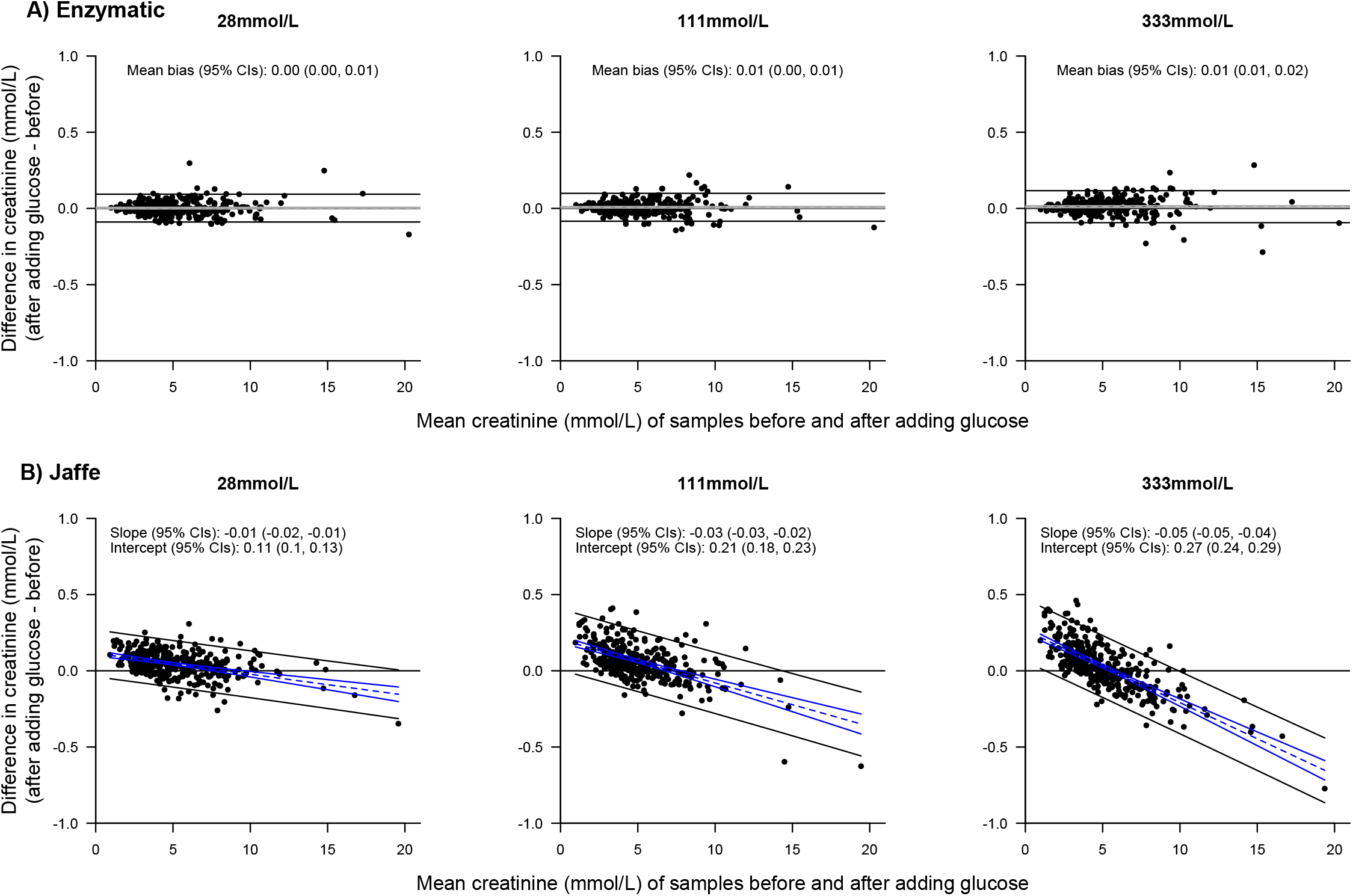
Bland-Altman plots for creatinine, by glucose concentration. Mean difference and 95% CIs are shown by dashed and solid grey lines. If slope is significantly different from 0, bias is instead shown by a regression line in blue. 95% limits of agreement are shown as solid black lines.

**Figure 2:**
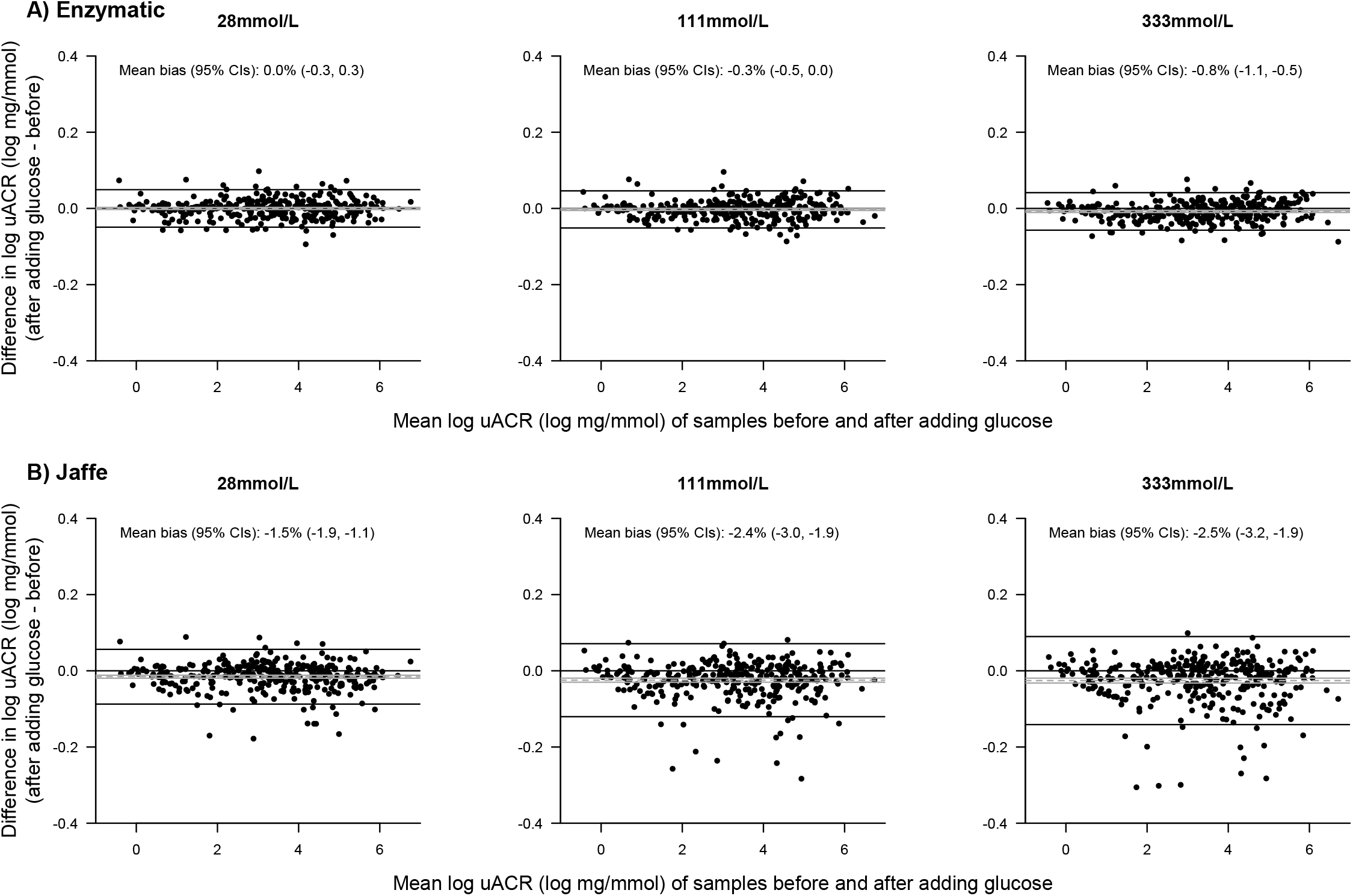
Bland-Altman plots for uACR, by glucose concentration. Mean bias and 95% CIs are shown by dashed and solid grey lines. 95% limits of agreement are shown as solid black lines. For log transformed variables, mean bias values have been back-transformed onto the original scale to give a percentage difference.

For the Jaffe creatinine method, the presence of glucose caused a bias which resulted in substantial overestimation of urinary creatinine at the lowest creatinine concentrations and a small underestimate at high urinary creatinine concentrations (Figure 1B). This bias was not importantly different across the range of levels of uACR (Figure S2). The net bias resulting from glucose interference was, on average, to underestimate uACR across the range of albumiuria studied. Figure 1B’s Bland-Altman plots show an increasingly steep regression line slope, indicating increasing bias with higher glucose concentrations. In this cohort, the presence of 28 mmol/L of glucose in the urine resulted in a -1.5% mean bias in uACR (95% confidence interval -1.9% to -1.1%) which increased to -2.5% (−3.2% to -1.9%) at a glucose concentration of 333 mmol/L (Figure 2B). Bias was largest at low creatinine concentrations (i.e. in dilute urine, Figure 3).

**Figure 3:**
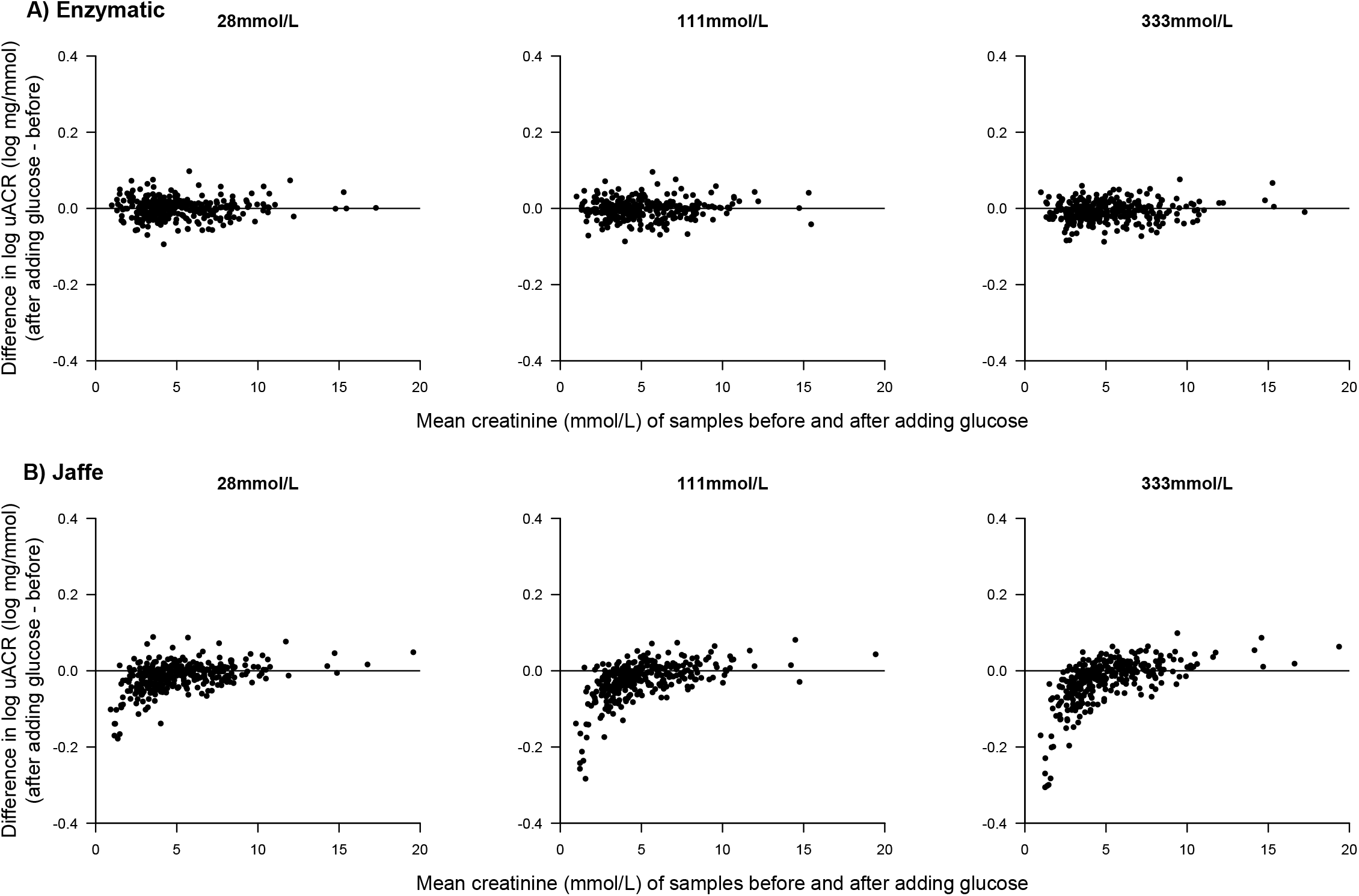
Association between mean creatinine and the difference in uACR before and after adding glucose, by assay method and glucose level.

### Illustrations of the impact of glucose interference on uACR (original scale)

Table 2 shows by how much uACR would be underestimated by a Jaffe assay in the presence of urinary glucose. In this particular cohort, interference from the highest level of glucose concentration led to 5.1% (17/333) and 1.8% of samples (6/333) having uACR underestimates of ≥10% and ≥20%, respectively. Table 3 provides absolute and percentage change in uACR for different hypothetical scenarios. Among the 4% (14/333) of participants with a urine creatinine <2.5 mmol/L, the presence of 28 mmol/L of urinary glucose caused a bias of -5.2% to -7.0% (depending on level of uACR). This bias increased to -10.1% to -13.0% at a concentration of 333 mmol/L. In comparison biases were all <1.0% for those with urinary creatinine of ≥5 mmol/L.

**Table 2:** Percentage of samples with reduction in uACR due to glucose spiking.

| Assay | Glucose concentration (mmol/L) | n (%) of samples with reduction in uACR due to glucose spiking alone |  |  |
| --- | --- | --- | --- | --- |
|  |  | ≥10% | ≥20% | ≥30% |
| Enzymatic | 28 | 0 | 0 | 0 |
|  | 111 | 0 | 0 | 0 |
|  | 333 | 0 | 0 | 0 |
| Jaffe | 28 | 7 (2%) | 0 | 0 |
|  | 111 | 12 (4%) | 4 (1%) | 0 |
|  | 333 | 17 (5%) | 6 (2%) | 0 |

uACR=urinary albumin-to-creatinine ratio, which is presented on its original scale.

**Table 3:** Absolute and percent change in uACR due to glucose interference of urine Jaffe assays by hypothetical levels of uACR.

| Glucose concentration (mmol/L) | Level of uACR (mg/mmol) | Urine creatinine concentration |  |  |  |  |  |
| --- | --- | --- | --- | --- | --- | --- | --- |
|  |  | <2.5 mmol/L |  | ≥2.5 to <5 mmol/L |  | ≥5 mmol/L |  |
|  |  | Absolute change (mg/mmol) | % | Absolute change (mg/mmol) | % | Absolute change (mg/mmol) | % |
| 28 | 3 | -0.21 | -7.0% | -0.01 | -0.3% | -0.02 | -0.7% |
| 28 | 30 | -1.55 | -5.2% | -0.48 | -1.6% | 0.04 | 0.1% |
| 28 | 300 | -17.39 | -5.8% | -6.29 | -2.1% | -0.99 | -0.3% |
| 111 | 3 | -0.28 | -9.3% | -0.05 | -1.7% | 0.02 | 0.7% |
| 111 | 30 | -2.86 | -9.5% | -0.84 | -2.8% | 0.06 | 0.2% |
| 111 | 300 | -31.22 | -10.4% | -9.61 | -3.2% | -0.24 | -0.1% |
| 333 | 3 | -0.32 | -10.7% | -0.03 | -1.0% | 0.01 | 0.3% |
| 333 | 30 | -3.04 | -10.1% | -0.92 | -3.1% | 0.23 | 0.8% |
| 333 | 300 | -39.11 | -13.0% | -11.61 | -3.9% | 2.71 | 0.9% |

uACR=urinary albumin-to-creatinine ratio, which is presented on its original scale. Percentage change in uACR is calculated as $e^b - 1$ , where b is the mean absolute bias in log uACR for the stated glucose concentration and absolute change in uACR is calculated by this proportional change to the hypothetical level of uACR. Mean creatinine concentration in the <2.5, ≥2.5 to <5 mmol/L, and ≥5 mmol/L groups were 1.8, 3.7 and 7.4 mmol/L respectively.

## Discussion

Jaffe assays are commonly used to measure creatinine and, the presence of glycosuria in the range expected to result from use of SGLT-2 inhibitors causes a biased underestimate of uACR when such an assay is used. This bias increases progressively with higher urinary glucose concentrations, and particularly affects dilute urine samples (urinary creatinine <2.5 mmol/L). Any underestimation of uACR by glucose interference creates a positive bias for any observed reduction in uACR in serial uACR measurements. In patients with CKD stage 3-4 in this study, high urinary glucose concentration resulted in a ∼10% underestimate of uACR among those with dilute urine. In contrast, enzymatic methods were almost unaffected.

Such a level of bias is arguably unacceptable if it alters decisions made by clinicians unaware of the interference. An overestimate of the reduction in albuminuria observed after starting an SGLT-2 inhibitor could, for example, result in a decision not to start other proven renal disease-modifying treatments, or make a patient ineligible for a treatment reserved for people with a certain level of albuminuria.^27^

This bias also has implications for analyses from historical and, design of future clinical trials. A 30% reduction in geometric mean uACR has been suggested as a meaningful and valid surrogate of treatments for progressive CKD.^28^ Although urinary glucose alone does not interfere with Jaffe assays sufficiently to result in this level of change, it could result in overestimated or misleading claims of beneficial effects on uACR. We were only able to identify type of urine assay used in one of eleven large placebo-controlled SGLT-2 inhibitor trials and none of four intensive versus standard glycaemic control trials (Supplementary Table 1).^29^

Given the increasing use of SGLT-2 inhibitors in clinical practice, we suggest uACR measured using Jaffe creatinine assays should be avoided. Alternatively alongside the results, assay type should be reported especially where resource limitations preclude the use of enzymatic methods. Enzymatic creatinine methods should be used in trials assessing effects of interventions on albuminuria when such interventions could modify glycosuria.

## Data Availability

All data produced in the present work are contained in the manuscript.

## Acknowledgements

We thank the participants, local clinical centre staff, and members of the steering and data monitoring committees. The UK HARP-III trial was designed, conducted, and analysed by the Medical Research Council Population Health Research Unit at the University of Oxford, which is part of the Clinical Trial Service Unit and Epidemiological Studies Unit (CTSU), Nuffield Department of Population Health. The University of Oxford was the sponsor for the trial, which was funded by a grant to the University of Oxford from Novartis (the manufacturer of sacubitril/valsartan). The funder had no involvement in the study conduct, analysis, or decision to submit this manuscript for publication. This study was supported by the Medical Research Council (MC_UU_00017/3) and the National Institute for Health Research Clinical Research Network. WGH was supported by a Medical Research Council Kidney Research UK Professor David Kerr Clinician Scientist Award (MR/R007764/1). Data used in this publication is available in line with the policy and procedures described at: https://www.ndph.ox.ac.uk/data-access. For further information, contact the corresponding author.

## Disclosures

CTSU has a staff policy of not accepting honoraria or other payments from the pharmaceutical industry, except for the reimbursement of costs to participate in scientific meetings (www.ctsu.ox.ac.uk). All authors report a grant to their institution from Boehringer Ingelheim to conduct the EMPA-KIDNEY trial.

## Author contributions

DC, WH & MH conceived the study. DC, TA, SC, SM, & MH designed the interference studies. RH, PJ, WH, MJL, & CB collected the data. DC & TA supervised the laboratory analyses. DC, PJ, WH and NS specified the statistical analyses which were performed by RS. DC, PJ, RS, MH, & WH drafted the manuscript which was reviewed and edits for content by all authors.

## Open access

For the purpose of open access, the author(s) has applied a Creative Commons Attribution (CC BY) licence to any Author Accepted Manuscript version arising.

## Notes

### Competing Interest Statement

The authors have declared no competing interest.

### Funding Statement

This study did not receive any funding.

### Author Declarations

Ethical (Nottingham Research Ethics Committee 2 [13/EM/0434]) and regulatory approvals were obtained before the enrollment of any study participants.

## References

1. Ginsberg JM, Chang BS, Matarese RA, Garella S. Use of single voided urine samples to estimate quantitative proteinuria. N Engl J Med. 1983; 309(25): 1543–6.

2. Stevens PE, Levin A, Kidney Disease: Improving Global Outcomes Chronic Kidney Disease Guideline Development Work Group M. Evaluation and management of chronic kidney disease: synopsis of the kidney disease: improving global outcomes 2012 clinical practice guideline. Ann Intern Med. 2013; 158(11): 825–30.

3. Inker LA, Levey AS, Pandya K, et al. Early change in proteinuria as a surrogate end point for kidney disease progression: an individual patient meta-analysis. Am J Kidney Dis. 2014; 64(1): 74–85.

4. Heerspink HJ, Kropelin TF, Hoekman J, de Zeeuw D, Reducing Albuminuria as Surrogate Endpoint C. Drug-Induced Reduction in Albuminuria Is Associated with Subsequent Renoprotection: A Meta-Analysis. J Am Soc Nephrol. 2015; 26(8): 2055–64.

5. Vallon V, Thomson SC. Targeting renal glucose reabsorption to treat hyperglycaemia: the pleiotropic effects of SGLT2 inhibition. Diabetologia. 2017; 60(2): 215–25.

6. Staplin N, Roddick AJ, Emberson J, et al. Net effects of sodium-glucose co-transporter-2 inhibition in different patient groups: a meta-analysis of large placebo-controlled randomized trials. EclinicalMedicine. 2021; 41: 101163.

7. Petrykiv SI, Laverman GD, de Zeeuw D, Heerspink HJL. The albuminuria-lowering response to dapagliflozin is variable and reproducible among individual patients. Diabetes Obes Metab. 2017; 19(10): 1363–70.

8. Petrykiv S, Sjostrom CD, Greasley PJ, Xu J, Persson F, Heerspink HJL. Differential Effects of Dapagliflozin on Cardiovascular Risk Factors at Varying Degrees of Renal Function. Clin J Am Soc Nephrol. 2017; 12(5): 751–9.

9. Cherney D, Lund SS, Perkins BA, et al. The effect of sodium glucose cotransporter 2 inhibition with empagliflozin on microalbuminuria and macroalbuminuria in patients with type 2 diabetes. Diabetologia. 2016; 59(9): 1860–70.

10. Barnett AH, Mithal A, Manassie J, et al. Efficacy and safety of empagliflozin added to existing antidiabetes treatment in patients with type 2 diabetes and chronic kidney disease: a randomised, double-blind, placebo-controlled trial. Lancet Diabetes Endocrinol. 2014; 2(5): 369–84.

11. Fioretto P, Stefansson BV, Johnsson E, Cain VA, Sjostrom CD. Dapagliflozin reduces albuminuria over 2 years in patients with type 2 diabetes mellitus and renal impairment. Diabetologia 2016; 59(9): 2036–9.

12. Jongs N, Greene T, Chertow GM, et al. Effect of dapagliflozin on urinary albumin excretion in patients with chronic kidney disease with and without type 2 diabetes: a prespecified analysis from the DAPA-CKD trial. Lancet Diabetes Endocrinol. 2021; 9(11): 755–66.

13. Cook JGH. Factors Influencing the Assay of Creatinine: Prepared for the Association of Clinical Biochemists’ Scientific and Technical Committee. Ann Clin Biochem. 1975; 12(1-6): 219–32.

14. Çuhadar S, Köseoğlu M, Çinpolat Y, Buğdaycı G, Usta M, Semerci T. The effect of extremely high glucose concentrations on 21 routine chemistry and thyroid Abbott assays: interference study. Biochem Med. 2016; 26(1): 53–60.

15. Küme T, Sağlam B, Ergon C, Sisman AR. Evaluation and comparison of Abbott Jaffe and enzymatic creatinine methods: Could the old method meet the new requirements? J Clin Lab Anal. 2018; 32(1):e22168.

16. UK NEQAS BQ. Urine Chemistries external quality assurance programme, Distribution 186. 2022 (accessed 22/05/2022 2022).

17. Wanner C, Inzucchi SE, Lachin JM, et al. Empagliflozin and Progression of Kidney Disease in Type 2 Diabetes. N Engl J Med. 2016; 375(4): 323–34.

18. Perkovic V, Jardine MJ, Neal B, et al. Canagliflozin and Renal Outcomes in Type 2 Diabetes and Nephropathy. N Engl J Med. 2019; 380(24): 2295–306.

19. Heerspink HJL, Stefánsson BV, Correa-Rotter R, et al. Dapagliflozin in Patients with Chronic Kidney Disease. N Engl J Med. 2020; 383(15): 1436–46.

20. Packer M, Anker SD, Butler J, et al. Cardiovascular and Renal Outcomes with Empagliflozin in Heart Failure. N Engl J Med. 2020; 383(15): 1413–24.

21. Anker SD, Butler J, Filippatos G, et al. Empagliflozin in Heart Failure with a Preserved Ejection Fraction. N Engl J Med. 2021; 385(16): 1451–61.

22. UK HARP-III Collaborative Group. Randomized multicentre pilot study of sacubitril/valsartan versus irbesartan in patients with chronic kidney disease: United Kingdom Heart and Renal Protection (HARP)-III-rationale, trial design and baseline data. Nephrol Dial Transplant. 2017; 32(12): 2043–51.

23. Herrington W, Illingworth N, Staplin N, et al. Effect of Processing Delay and Storage Conditions on Urine Albumin-to-Creatinine Ratio. Clin J Am Soc Nephrol. 2016; 11(10): 1794–801.

24. Chapman DP, Gooding KM, McDonald TJ, Shore AC. Stability of urinary albumin and creatinine after 12 months storage at -20 degrees C and -80 degrees C. Pract Lab Med. 2019; 15: e00120.

25. Kim SR, Lee YH, Kang ES, Cha BS, Lee BW. The Relationship between Increases in Morning Spot Urinary Glucose Excretion and Decreases in HbA1C in Patients with Type 2 Diabetes After Taking an SGLT2 Inhibitor: A Retrospective, Longitudinal Study. Diabetes Ther. 2017; 8(3): 601–9.

26. Martin Bland J, Altman D. Statistical methods for assessing agreement between two methods of clinical measurement. Lancet. 1986; 327(8476): 307–10.

27. NICE. Dapagliflozin for treating chronic kidney disease. 2022. https://www.nice.org.uk/guidance/ta775/chapter/1-Recommendations (accessed 22/08/2022 2022).

28. Heerspink HJL, Greene T, Tighiouart H, et al. Change in albuminuria as a surrogate endpoint for progression of kidney disease: a meta-analysis of treatment effects in randomised clinical trials. Lancet Diabetes Endocrinol. 2019; 7(2): 128–39.

29. 29. Zoungas S, Arima H, Gerstein HC, et al. Effects of intensive glucose control on microvascular outcomes in patients with type 2 diabetes: a meta-analysis of individual participant data from randomised controlled trials. Lancet Diabetes Endocrinol. 2017; 5(6): 431–7.

