## Supplementary Figures 1 & 2 for "Interference of urinary albumin-to-creatinine ratio measurement by glycosuria: clinical implications when using SGLT-2 inhibitors"

### Figure S1: Bland-Altman plots for albumin, by glucose concentration

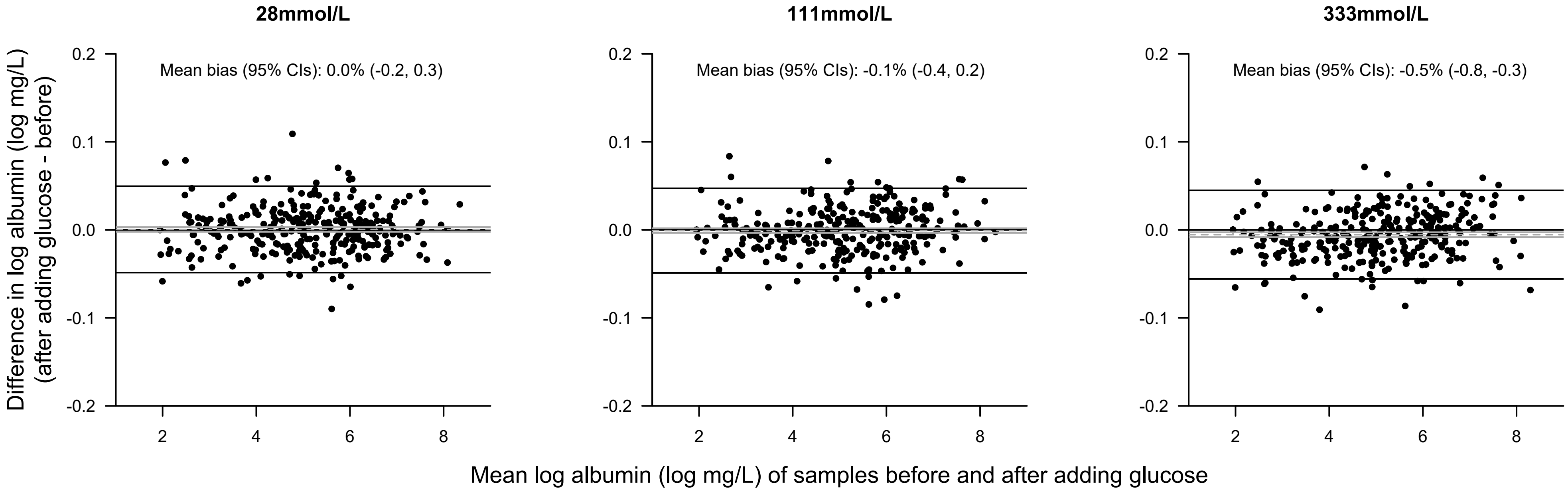


Mean bias and 95% CIs are shown by dashed and solid grey lines. 95% limits of agreement are shown as solid black lines.
For log transformed variables, mean bias values have been back-transformed onto the original scale to give a percentage difference.

#### Figure S2: Association between creatinine and the difference in uACR before and after adding glucose to a Jaffe assay, by glucose level and uACR at randomization.

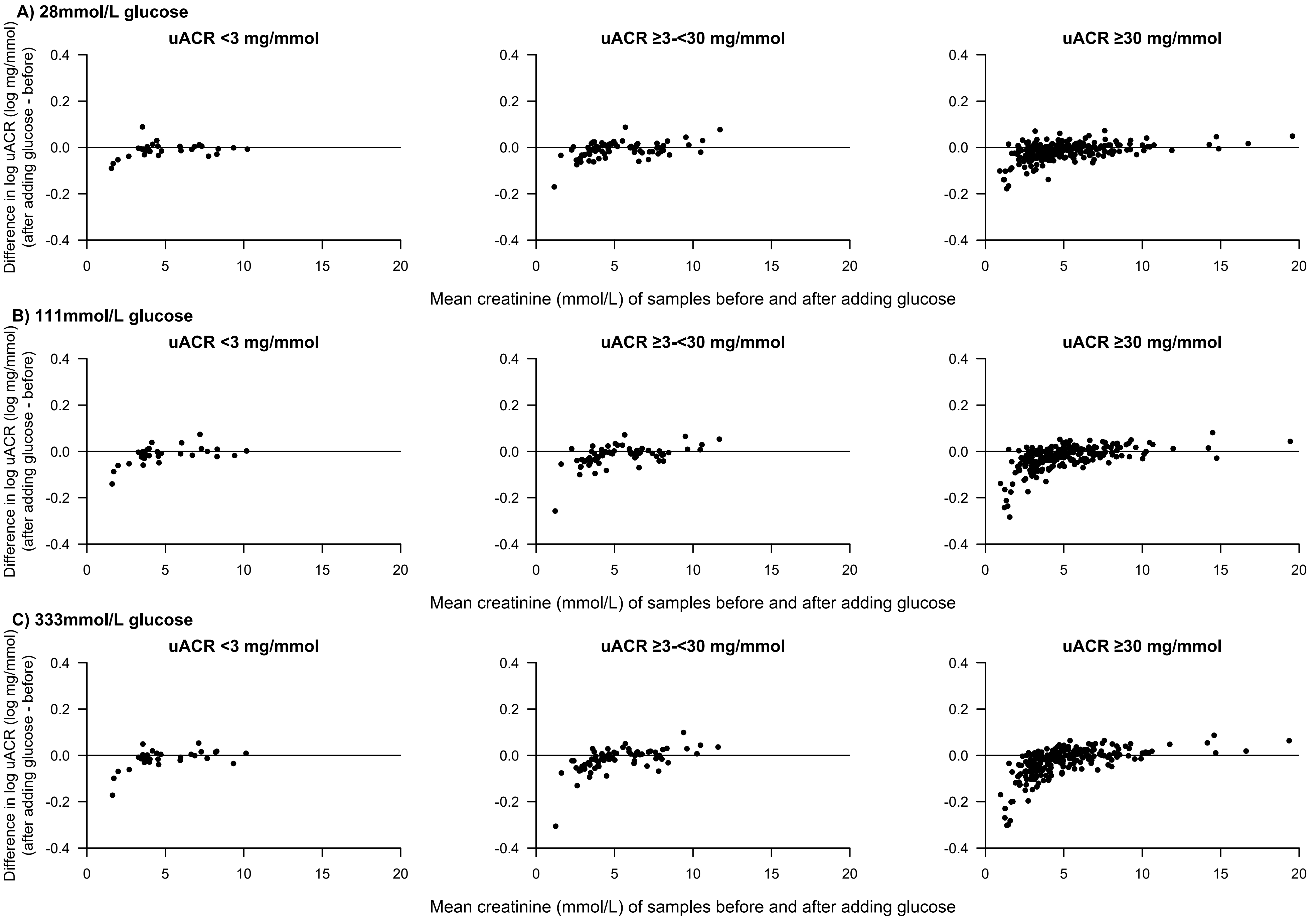
