## Supplementary Table 1 for "Interference of urinary albumin-to-creatinine ratio measurement by glycosuria: clinical implications when using SGLT-2 inhibitors"

### Supplementary Table 1: Methods of albuminuria measurement in other clinical trials

| Trial acronym | uACR measurement method recorded |
|---|---|
| **SGLT2-inhibitor trials** | |
| DECLARE-TIMI 58[1,2] | No |
| CANVAS Program[3,4] | UACR was measured every 26 weeks in CANVAS-R, and at week 12 and then annually in CANVAS. Serum creatinine measurement with estimation of GFR was undertaken in a central laboratory using the Jaffe method with rate blanking |
| VERTIS CV[5] | No |
| EMPA-REG OUTCOME[6,7] | No |
| DAPA-HF[8] | No |
| EMPEROR-REDUCED[9] | No |
| EMPEROR-PRESERVED[10] | No |
| CREDENCE[3] | No |
| SOLOIST-WHF[11] | UACR not tested. |
| SCORED[12] | No |
| DAPA-CKD[13] | No |
| **Other trials** | |
| ACCORD[14] | No |
| UKPDS[15,16] | From 1988, urine albumin measured by immunoturbidimetric method |
| ADVANCE[17] | No |
| VADT[18] | No |

**References**


1. Wiviott SD, Raz I, Bonaca MP, et al. Dapagliflozin and Cardiovascular Outcomes in Type 2 Diabetes. N Engl J Med. 2019;380:347-57.
2. Mosenzon O, Wiviott SD, Cahn A, et al. Effects of dapagliflozin on development and progression of kidney disease in patients with type 2 diabetes: an analysis from the DECLARE-TIMI 58 randomised trial. Lancet Diabetes Endocrinol. 2019;7:606-17.
3. Perkovic V, Jardine MJ, Neal B, et al. Canagliflozin and Renal Outcomes in Type 2 Diabetes and Nephropathy. N Engl J Med. 2019;380:2295-306.
4. Perkovic V, de Zeeuw D, Mahaffey KW, et al. Canagliflozin and renal outcomes in type 2 diabetes: results from the CANVAS Program randomised clinical trials. Lancet Diabetes Endocrinol. 2018;6:691-704.
5. Cannon CP, Pratley R, Dagogo-Jack S, et al. Cardiovascular Outcomes with Ertugliflozin in Type 2 Diabetes. N Engl J Med. 2020;383:1425-35.
6. Wanner C, Inzucchi SE, Lachin JM, et al. Empagliflozin and Progression of Kidney Disease in Type 2 Diabetes N Engl J Med. 2016;375:323-34.
7. Cherney DZI, Zinman B, Inzucchi SE, et al. Effects of empagliflozin on the urinary albumin-to-creatinine ratio in patients with type 2 diabetes and established cardiovascular disease: an exploratory analysis from the EMPA-REG OUTCOME randomised, placebo-controlled trial. Lancet Diabetes Endocrinol. 2017;5:610-21.
8. McMurray JJV, Solomon SD, Inzucchi SE, et al. Dapagliflozin in Patients with Heart Failure and Reduced Ejection Fraction. N Engl J Med. 2019;381:1995-2008.
9. Packer M, Anker SD, Butler J, et al. Cardiovascular and Renal Outcomes with Empagliflozin in Heart Failure. N Engl J Med. 2020;383:1413-24.
10. Anker SD, Butler J, Filippatos G, et al. Empagliflozin in Heart Failure with a Preserved Ejection Fraction. N Engl J Med. 2021;385:1451-61.
11. Bhatt DL, Szarek M, Steg PG, et al. Sotagliflozin in Patients with Diabetes and Recent Worsening Heart Failure. N Engl J Med. 2020;384:117-28.
12. Bhatt DL, Szarek M, Pitt B, et al. Sotagliflozin in Patients with Diabetes and Chronic Kidney Disease. N Engl J Med. 2020;384:129-39.
13. Heerspink HJL, Stefánsson BV, Correa-Rotter R, et al. Dapagliflozin in Patients with Chronic Kidney Disease. N Engl J Med. 2020;383:1436-46.
14. Group AS, Cushman WC, Evans GW, et al. Effects of intensive blood-pressure control in type 2 diabetes mellitus. N Engl J Med. 2010;362:1575-85.
15. UK Prospective Diabetes Study Group. Tight blood pressure control and risk of macrovascular and microvascular complications in type 2 diabetes: UKPDS 38. UK Prospective Diabetes Study Group. BMJ (Clinical research ed). 1998;317:703-13.
16. UK Prospective Diabetes Study Group. UK Prospective Diabetes Study (UKPDS). VIII. Study design, progress and performance. Diabetologia. 1991;34:877-90.
17. The ADVANCE Collaborative Group. Intensive Blood Glucose Control and Vascular Outcomes in Patients with Type 2 Diabetes. N Engl J Med. 2008;358:2560-72.
18. Duckworth W, Abraira C, Moritz T, et al. Glucose Control and Vascular Complications in Veterans with Type 2 Diabetes. N Engl J Med. 2009;360:129-39.
